# Acceptance and Attitudes Toward COVID-19 Vaccines: A Cross-Sectional Study from Jordan

**DOI:** 10.1101/2020.12.22.20248676

**Authors:** Tamam El-Elimat, Mahmoud M. AbuAlSamen, Basima A. Almomani, Nour A. Al-Sawalha, Feras Q. Alali

**Affiliations:** Department of Medicinal Chemistry and Pharmacognosy, Faculty of Pharmacy, Jordan University of Science and Technology, Irbid 22110, Jordan; Department of Family and Community Medicine, Faculty of Medicine, University of Jordan, Amman 11942, Jordan; Department of Clinical Pharmacy, Faculty of Pharmacy, Jordan University of Science and Technology, Irbid 22110, Jordan; Faculty of Pharmacy, QU Health, Qatar University, Doha 2713, Qatar; Biomedical and Pharmaceutical Research Unit, QU Health, Qatar University, Doha 2713, Qatar

**Keywords:** COVID-19, Attitudes, Vaccines, acceptance, public, Jordan

## Abstract

**Background:** Vaccines are effective interventions that can reduce the high burden of diseases globally. However, public vaccine hesitancy is a pressing problem for public health authorities. With the availability of COVID-19 vaccines, little information is available on the public acceptability and attitudes towards the COVID-19 vaccines in Jordan. This study aimed to investigate the acceptability of COVID-19 vaccines and its predictors in addition to the attitudes towards these vaccines among public in Jordan.

**Methods:** An online, cross-sectional, and self-administered questionnaire was instrumentalized to survey adult participants from Jordan on the acceptability of COVID-19 vaccines. Logistic regression analysis was used to find the predictors of COVID-19 vaccines’ acceptability.

**Results:** A total of 3,100 participants completed the survey. The public acceptability of COVID-19 vaccines was fairly low (37.4%) in Jordan. Males (OR=2.488, 95CI%=1.834–3.375, *p*<.001) and those who took the seasonal influenza vaccine (OR=2.036, 95CI%=1.306–3.174, *p*=.002) were more likely to accept Covid-19 vaccines. Similarly, participants who believed that vaccines are generally safe (OR=9.258, 95CI%=6.020–14.237, *p*<.001) and those who were willing to pay for vaccines (OR=19.223, 95CI%=13.665–27.042, *p*<.001), once available, were more likely to accept the COVID-19 vaccines. However, those above 35 years old (OR=0.376, 95CI%=0.233-0.607, *p*<.001) and employed participants (OR=0.542, 95CI%=0.405-0.725, *p*<.001) were less likely to accept the COVID-19 vaccines. Moreover, participants who believed that there was a conspiracy behind COVID-19 (OR=0.502, 95CI%=0.356- 0.709, *p*<.001) and those who do not trust any source of information on COVID-19 vaccines (OR=0.271, 95CI%=0.183 – 0.400, *p*<.001), were less likely to have acceptance towards them. The most trusted sources of information on COVID-19 vaccines were healthcare providers.

**Conclusion:** Systematic interventions are required by public health authorities to reduce the levels of vaccines’ hesitancy and improve their acceptance. We believe these results and specifically the low rate of acceptability is alarming to Jordanian health authorities and should stir further studies on the root causes and the need of awareness campaigns. These interventions should take the form of reviving the trust in national health authorities and structured awareness campaigns that offer transparent information about the safety and efficacy of the vaccines and the technology that was utilized in their production.

## Introduction

Severe acute respiratory syndrome coronavirus 2 (SARS-CoV-2) is the causative virus for the coronavirus disease 2019 (COVID-19) ongoing pandemic [1-4]. SARS-CoV-2 first emerged in late 2019 in Wuhan (Hubei, China) and hastily become a global threat affecting 220 countries [1, 2]. As of 19 December, the COVID-19 pandemic has resulted in more than 74.2 M cases and more than 1.6 M deaths worldwide [1]. The pandemic has resulted in a devastating impact worldwide, which prompted the need for mitigation policies to contain the pandemic [5]. The ground strategy followed by most countries around the world was to reduce the transmissibility of the disease, often by non-pharmaceutical interventions (NPIs), including enforcing masks policy, hands sanitization, social distancing, travel restrictions, schools’ closures, and partial or complete lockdowns. [6]. So far, NPIs were able to slow down the progression of the disease, but the most promising strategy to confine the pandemic and providing hope to reduce the mortality and morbidity rates remains within the capacity of medical technology. Such medical technology includes effective, safe, and affordable antiviral agents and vaccines. As of December 2020, no antiviral drugs have been approved that were specifically developed against SARS-CoV-2 [7]. The US Food and Drug Administration (FDA) has granted remdesivir an Emergency Use Authorization for severely ill hospitalized patients with COVID-19 [8, 9]. However, the WHO recommended against its use in November 2020 [10].

Vaccines are one of the most reliable and cost-effective public health interventions ever implemented that are saving millions of lives each year [11-13]. Following the deciphering of the genome sequence of SARS-CoV-2 in early 2020 [3] and the declaration of the pandemic by WHO in March 2020 [4], scientists and pharmaceutical companies are racing against time in efforts to develop vaccines [14, 15]. As of December 19, 2020, at least 85 vaccines are in preclinical development in animals and 63 are in clinical development in humans, of which 43 in phase I, 20 in phase II, 18 in phase III, 6 have been approved for early or limited use, 2 have been approved for full use, and one vaccine has been abandoned [14]. Pfizer-BioNTech’s (BNT162b2) and Moderna (mRNA-1273) mRNA vaccines have been approved for emergency use in the US [14].

With the uplifting news about SARS-CoV-2 vaccines approval, optimism is raising to see an end to the pandemic through herd immunity [16, 17]. The threshold for SARS-CoV-2 herd immunity is estimated to range between 50% and 67% [16]. One major obstacle facing the achievement of such a goal is believed to be vaccine hesitancy and skepticism among the population worldwide [15, 18, 19]. Vaccine hesitancy was defined by the WHO Strategic Advisory Group of Experts (SAGE) as “*delay in acceptance or refusal of vaccination despite availability of vaccination services*” [20]. Vaccine acceptability is determined by three factors: confidence, convenience, and complacency [29]. Confidence is defined as the trust in the safety and effectiveness of the vaccine, trust in the delivery system as the healthcare system, and the trust in the policymakers [28]. Many people have doubts about vaccine safety, and this is going to be a major challenge to be resolved by health care providers, policymakers, community leaders, and governments to increase the widespread acceptance of the vaccines [15, 18, 19]. Moreover, vaccination convenience refers to the relative ease of access to the vaccine that includes physical availability, affordability, and accessibility [20]. Vaccine complacency is associated with a low realized risk of the vaccine-preventable disease and hence more negative attitudes towards the vaccines [28]. Such skepticism was demonstrated in a poll that was conducted in the US, where 50% of the Americans said they are willing to take the vaccine, 30% are unsure, while 20% are refusing the vaccine [21]. In another survey of adult Americans, 58% intended to be vaccinated, 32% were not sure, and 11% did not intend to be vaccinated [22]. However, one more study reported 67% of the Americans would accept a COVID-19 vaccine if it is recommended to them, although there were significant demographic differences in vaccine acceptance [23].

Jordan, with a population of 10 M, has one of the highest per-capita rates of COVID-19 infection in the world [1, 24]. As of December 2020, Jordan has reported more than 271,000 COVID-19 confirmed cases and over 3,500 deaths [1, 24]. This drastic increase, which started in September 2020 was embarked upon after a few months of the control of the pandemic in the country by implementing a very strict preventive lockdown that had painful societal and economic impacts [25]. The Jordanian government has announced that the first doses of Pfizer-BioNTech’s vaccine are going to be available by the end of January 2021 [26]. Enough vaccine doses to achieve the herd immunity threshold will not be available till 2022. Hence, it is crucial to explore the acceptance of COVID-19 vaccines and its predictors as well as the attitudes towards COVID-19 vaccines among Jordanian population. The results of current study could assist the policymakers to undertake proactive campaigns and well-designed strategies by highlighting the importance of vaccination to the community and encouraging vaccine uptake and acceptance, especially by vulnerable patients to stop further deaths and to confine the spread of the pandemic.

## Materials and Methods

### Study Design

A cross-sectional survey-based study was conducted in November 2020. A convenient sample approach was adopted in this study where people from the different Jordanian regions (Northern, Central, and Southern) were invited to participate. Amid the global pandemic, researchers utilized social media platforms to collect data. In this study, online social media platforms (Facebook, WhatsApp) were used to recruit participants. Participants were encouraged to pass on the questionnaire to their contacts or acquaintances. The main outcome of the study was the public acceptance of the COVID-19 vaccine. The study was approved by the Institutional Review Board at Jordan University of Science and Technology (protocol number 816/2020).

### Instrument Development and Measures

The questionnaire used in this study was developed based on literature review and discussion within the research team. The questionnaire was reviewed by experts in survey research for face validity. The questionnaire was structured into 4 sections. A pilot sample (n=26) was used to improve the wording and clarity of expression of the survey items. Data from the pilot sample was not used in any further analysis. The final version of the questionnaire required an estimated time of 5-10 minutes to complete.

### Sociodemographic Characteristics and Medical History

The sociodemographic characteristics of the participants were obtained as described below. Data collected were age, gender, marital status, smoking, employment status, academic level, and medical insurance. Additionally, participants were asked to report their history with chronic conditions and whether they took a seasonal flu vaccine this year or not.

### COVID-19 Pandemic Related Information

Participants were asked to indicate if they were infected with COVID-19 or knew anyone who was infected with confirmation of diagnosis using standard laboratory testing protocols. Another question item was dedicated to surveying participants who believe they may have contracted the virus but without a confirming test.

Participants were asked to indicate their most trusted sources when seeking knowledge of COVID-19 vaccines. Besides, participants were asked about their concerns during the COVID-19 pandemic.

### Acceptance of COVID-19 Vaccines

Participants were asked whether they accept to receive COVID-19 vaccines when they are approved and available in Jordan with 3 response levels (non-acceptance, neutral, acceptance). Variables that were investigated as potential predictors of COVID-19 vaccines acceptance were: age, gender, marital status, having children, academic area, employment, smoking status, whether the person received a seasonal flu vaccine this year, stating that vaccines are safe, concerned that there is a conspiracy behind COVID-19 pandemic, not having any trust in any source of information on vaccines, and willingness to pay for COVID-19 vaccines.

### Attitudes Toward COVID-19 Vaccines

The attitudes towards COVID-19 vaccines’ section consists of 6 statements with a 5-point Likert scale (5=strongly agree, 4=agree, 3=neutral, 2=disagree, 1=strongly disagree), with questions about hesitancy and concerns regarding COVID-19 vaccines.

### Statistical Analysis

Categorical variables were presented as numbers and percentages, while continuous variables were presented as median [interquartile range]. The univariate analysis was performed using an independent Mann–Whitney U test for continuous variables and Chi-square test for categorical variables as appropriate. For analysis, responses to the attitudes section were combined. For example, both responses “strongly agree” and “agree” were combined in one category and both responses “strongly disagree” and “disagree” in one category.

The main outcome of the study was the public acceptance of COVID-19 vaccines. To determine the factors that affect the acceptance of the Jordanian population to receive COVID-19 vaccines, both multinomial and binary logistic regressions were performed. At first, potential predictors for COVID-19 vaccines were screened using univariable analysis, and variables with *p*<.05 were considered in both multinomial and binary logistic regression. When the multinomial logistic regression was conducted, the acceptance outcome was trichotomized as (non-acceptance, neutral, and acceptance). For a simpler interpretation of the analysis, the participants who answered ‘neutral’ were then removed and a binary logistic regression was performed. In the binary logistic regression model, the participants were dichotomized as acceptable or not acceptable. In both models, the odds ratio (OR) values and their 95% confidence intervals (95% CI) were calculated. A *p*-value of less than .05 was considered statistically significant. The analysis was carried out using the Statistical Package for Social Sciences (SPSS Inc., Chicago, IL) version 23.

## Results

### Demographics

A total of 3,100 participants were enrolled in the current study. The median age of participants was 29 years old and more than half of them (67.4%) were females. Half of the respondents (49.8%) were married and had kids (46.1%). About 70% had an undergraduate degree and more than half (53.8%) with health-related educational backgrounds. Besides, 46.4% of the participants were employed and only 13.4% had chronic diseases. Detailed demographics are presented in Table 1.

**Table 1.**
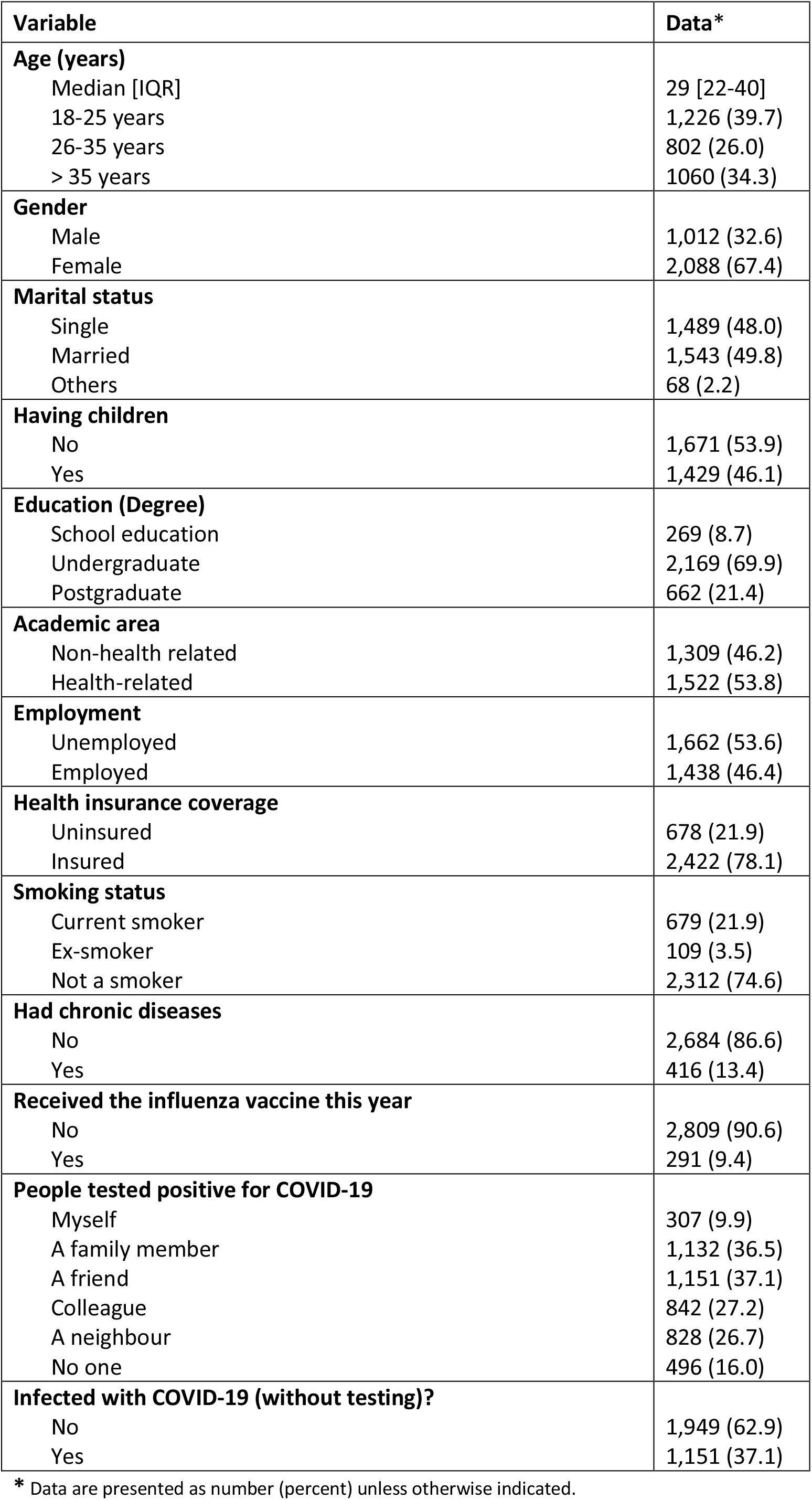
Demographic details of study participants (n=3,100)

**Table 2.** Attitudes toward COVID-19 vaccines

| Attitudes | Strongly agree/agree<br>N (%) | Neutral<br>N (%) | Strongly disagree/disagree<br>N (%) |
| --- | --- | --- | --- |
| It is important to get a vaccine to protect the people from COVID-19. | 2,062 (66.5) | 609 (19.6) | 429 (13.8) |
| Pharmaceutical companies are going to develop safe and effective COVID-19 vaccines. | 1,819 (58.7) | 825 (26.6) | 456 (14.7) |
| COVID-19 vaccines made in Europe or America are safer than those made in other world countries. | 954 (30.8) | 1,160 (37.4) | 986 (31.8) |
| Side effects will prevent me from taking a vaccine for the prevention of COVID-19. | 1,538 (49.6) | 748 (24.1) | 814 (26.3) |
| Most people will refuse to take the COVID-19 vaccine once licensed in Jordan. | 1,528 (49.3) | 791 (25.5) | 781 (25.2) |
| The government will make the vaccine available for all citizens for free? | 1,122 (36.2) | 837 (27.0) | 1,141 (36.8) |

Less than 10% of the participants received the influenza vaccine this year. About 10% of the respondents reported that they had tested positive for COVID-19. However, about one-third of the participants (37.1%) stated that they might have been infected with COVID-19, but they did not confirm it by any laboratory testing.

As shown in Figure 1, almost half (45.4%) of the participants trusted healthcare providers as a source of information about COVID-19 vaccines. About 30%, 17%, and 16% of the participants trusted pharmaceutical companies, the internet, and media as sources of information, respectively. However, 18.1% of the participants did not trust any source.

**Figure 1.**
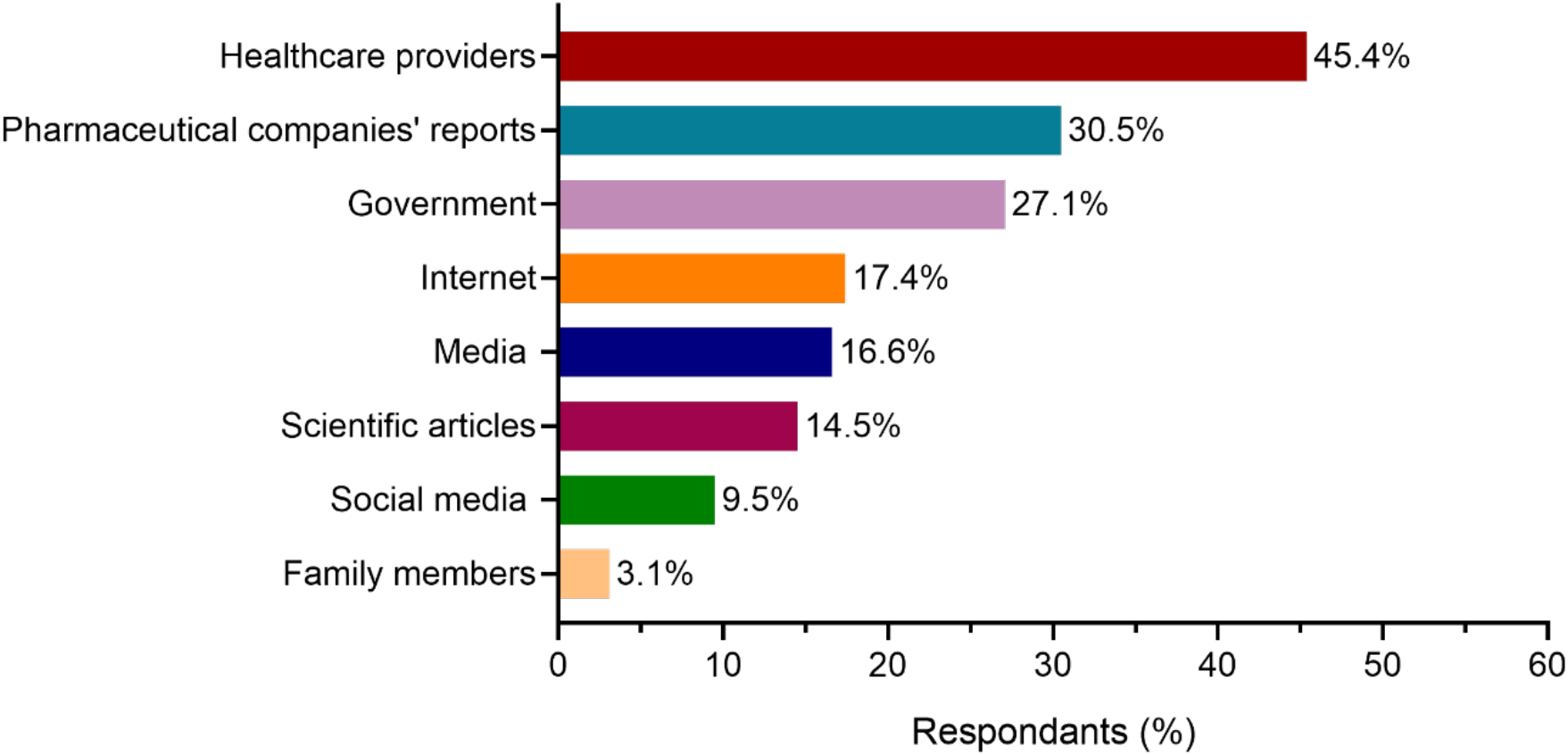
Most-trusted information sources about COVID-19 vaccines.

The participants were concerned about different issues during the COVID-19 pandemic (Figure 2). The most-reported concern by participants was a fear of family members being infected with COVID-19 (73.1%), which is higher than a concern about themselves being infected (27.3%). Almost one-third of the participants (30%) were concerned about death and 17.5% about the unavailability of vaccines. Approximately, a tenth of the participants was not worried about any issue.

**Figure 2.**
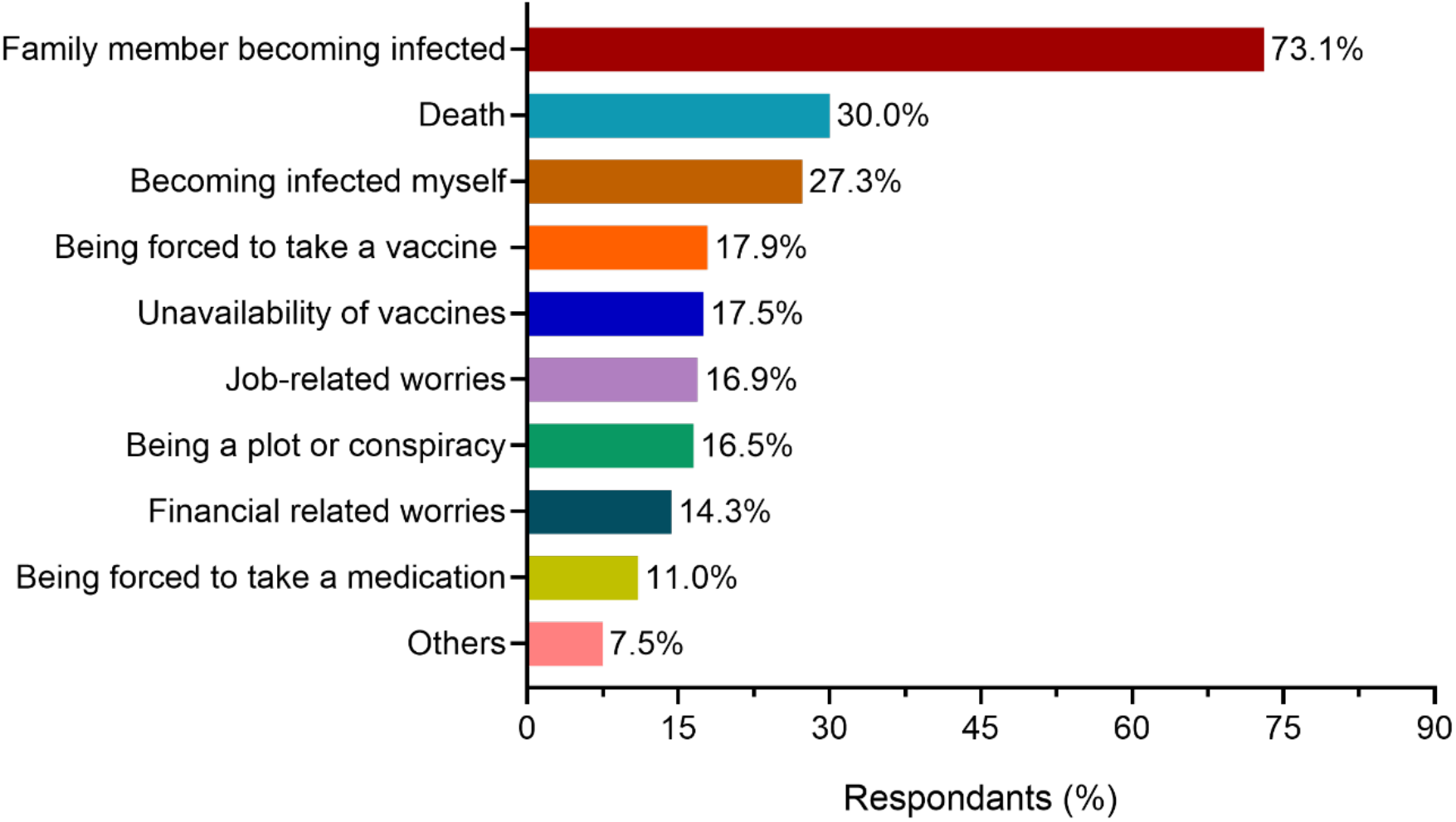
Jordan population worries during the COVID-19 pandemic.

### Acceptance for COVID-19 Vaccines

In the present study, 37.4% of the public were acceptable, 36.3% were not acceptable and 26.3% were neutral to receive COVID-19 vaccines. As shown in Table 3, the results of multivariate analysis (binary logistic regression) identified the independent factors that predicted the level of acceptance. The result indicated that the older age groups (>35 years old) were less likely to accept for COVID-19 vaccines compared to younger age groups (OR=0.376, 95CI%=0.233-0.607, *p*<.001). In addition, employed participants (OR=0.542, 95CI%=0.405-0.725, *p*<.001) were less likely to accept COVID-19 vaccines compared to unemployed participants. Furthermore, participants who believed that the COVID-19 pandemic is a conspiracy (OR=0.502, 95CI%=0.356-0.709, *p*<.001) and those who did not trust any information (OR=0.271, 95CI%=0.183 – 0.400, *p*<.001) were less acceptable for the vaccine.

**Table 3.** Predictors of acceptance for COVID-19 vaccines

| **Factors** | **OR** | **95% CI** | **P-value** |
| --- | --- | --- | --- |
| **Age** |  |  |  |
| 18-25 years | Ref |  |  |
| 26-35 years | 0.744 | 0.501–1.105 | 0.143 |
| >35 years | 0.376 | 0.233–0.607 | $p<.001$ |
| **Gender** |  |  |  |
| Female | Ref | | $p<.001$ |
| Male | 2.488 | 1.834–3.375 |  |
| **Employment** |  |  |  |
| Unemployed | Ref | | $p<.001$ |
| Employed | 0.542 | 0.405–0.725 |  |
| **Received the influenza vaccine this year** |  |  |  |
| No | Ref | | $p=.002$ |
| Yes | 2.036 | 1.306–3.174 |  |
| **Vaccines are safe** |  |  |  |
| No | Ref | | $p<.001$ |
| Not sure | 3.502 | 2.178–5.631 | $p<.001$ |
| Yes | 9.258 | 6.020–14.237 | $p<.001$ |
| **Concerned that COVID-19 pandemic is a conspiracy** |  |  |  |
| No | Ref | | $p<.001$ |
| Yes | 0.502 | 0.356–0.709 |  |
| **Not trust any information** |  |  |  |
| No | Ref | | $p<.001$ |
| Yes | 0.271 | 0.183–0.400 |  |
| **Willingness to pay for COVID-19 vaccines** |  |  |  |
| No | Ref | | $p<.001$ |
| Not sure | 3.895 | 2.984–5.084 | $p<.001$ |
| Yes | 19.223 | 13.665–27.042 | $p<.001$ |

CI: confidence interval; OR: odds ratio

On the other hand, males were more likely to have acceptance for COVID-19 vaccines (OR=2.488, 95CI%=1.834–3.375, *p*<.001) compared to females. In addition, participants who took the influenza vaccine this year were more likely to accept COVID-19 vaccines compared to those who did not take the influenza vaccine (OR=2.036, 95CI%=1.306–3.174, *p*=.002). Furthermore, participants who stated that vaccines are safe in general were 9 times more likely to accept taking COVID-19 vaccines compared to those who stated that vaccines are not safe (OR=9.258, 95CI%=6.020–14.237, *p*<.001). Moreover, participants who expressed their willingness to pay for COVID-19 vaccines were 19 times more likely to accept taking them compared to those who did not show their willingness to pay (OR=19.223, 95CI%=13.665–27.042, *p*<.001).

Similarly, results from multinomial logistic regression including the ‘neutral’ group, suggested that the predictors that make participants more likely to be in the acceptance group compared to the neutral group were being male (OR=1.522, 95% CI= 1.226–1.890, *p*<.0001) and among the age group of 18-25 years (OR=1.496, 95% CI= 1.083–2.067, *p*=.015). Additionally, people who did not believe that there is a conspiracy behind COVID-19 were also 1.4 times more likely to be in the acceptance group (OR=1.392, 95% CI= 1.022–1.898, *p*=.036). However, predictors that make people less likely to be in the acceptance group compared to the neutral group were people who did not take the seasonal flu vaccine this year (OR=0.675, 95% CI= 0.485–0.941, *p*=.02), those who believe that vaccines are unsafe (OR= 0.333, 95% CI= 0.223–0.498, p<.0001), and those who are not willing to pay for the vaccine once available (OR=0.321, 95% CI= 0.258–0.399, *p*<.0001).

### Attitudes Toward COVID-19 Vaccines

Almost two-thirds (66.5%) of the participants were strongly agreed/agreed that it is important to get a vaccine to protect people from COVID-19. Besides, less than 60% of the participants agreed that pharmaceutical companies will be able to develop safe and effective COVID-19 vaccines. Moreover, about half of the respondents (49.6%) reported that side effects will prevent them from taking a COVID-19 vaccine and that 49.3% will refuse to take COVID-19 vaccines once licensed. Importantly, around a quarter of all respondents were neutral regarding most attitudes as shown in Table 2.

## Discussion

This study sought to examine, for the first time, the Jordanians’ population acceptance of COVID-19 vaccines. Vaccine hesitancy could threaten the efficiency of COVID-19 vaccines once they become commercially available worldwide [27]. Compared to reports from studies conducted on public acceptance and willingness to receive the COVID-19 vaccines worldwide, Jordan stands among the lowest countries as 37.4% of our public were acceptable. A study based on a sample from 19 countries involving 13,426 participants showed that the global acceptance of COVID-19 vaccines ranges between as low as 54.8% from Russia to as high as 88.6% from China [28]. Moreover, most western countries report relatively higher public acceptance (59-75%) [28]. Similarly, Saudi Arabia, a country with similar demographic distributions as Jordan, reported a higher acceptance level (64.7%) [29]. The acceptability level of vaccinations in Jordan was often lower than global averages, including seasonal influenza vaccines in 2016-2018 [30, 31].

In the current study, predictors of COVID-19 vaccines from both multinomial and binary logistic regression were similar. Younger participants were more likely to accept COVID-19 vaccines in the current study, contrary to studies reporting higher acceptance among older age groups [23, 28, 29]. This can be explained by the different age distribution among countries, given Jordan as a country with mostly a young population and high literacy levels. Another reason could be related to the nature of the study design, this study could have been more biased towards the young, as the elderly are less likely to engage with online-based surveys. Moreover, there are contrasting reports of gender effects in the literature, wherein some males were more likely to accept the vaccine [23], compared to others reporting higher acceptance among females [28, 29]. In our study, Jordanian males were more likely to take the vaccine, in agreement with studies reported elsewhere [23]. Interestingly, Jordanian males were more likely to participate in COVID-19 vaccine clinical trials compared to females in 2020 [32]. Moreover, employed participants were less likely to accept taking the COVID-19 vaccines, in a contrasting result to available studies in the literature suggesting that employed individuals were more likely to accept COVID-19 vaccines. In our study, the employed participants were older than the unemployed ones, which were found to be less acceptable to get COVID-19 vaccines.

The low acceptance level of COVID-19 vaccines among Jordanians can be attributed to multi factors, some of which are shared with the wide global community. There is an evident uncertainty clouding the COVID-19 vaccines. Firstly, the new mRNA-based vaccines as a novel technology could be received with some skepticism since no prior experience or successes with such approach have been reported in the past. Also, the speed of vaccine development and registration in less than a year may have mediated a role in lowering the acceptance level. The current study revealed that half of the participants had safety concerns about the vaccine once it being available as indicated by their concerns about related side effects. This is consistent with Pogue and colleagues finding where the majority of participants (∼63%) in the USA stated that they were worried about the side effects of the COVID-19 vaccines [33]. Also, most of the participants (66.5%) in the current study stated that receiving the vaccine is important to protect against COVID-19. However, almost half of them (49%) agreed that most people would refuse to take the vaccine. This discrepancy could be due to their concerns about the vaccine’s side effects. Another global phenomenon that negatively contributed to such a low level is the numerous campaigns launched by anti-vaccinationists fueled by the new technology and short span of vaccine development. Such campaigns on social media with fabricated, false, and sometimes misleading Arabic translations feed the conspiracy beliefs of some people. Our results supported such perceived viewpoints, where those who did not believe in a conspiracy behind COVID-19 were more likely to accept COVID-19 vaccines. Some factors that are specific to the country and the region could also play a role in this. For example, there is a sector of the public who had their trust shaken in local Jordanian authorities and/or disapprove the overall handling of the pandemic. Some people expressed their frustration as many decisions were unwelcomed, disproportional with the pandemic status, not justified or backed with science. During August-October 2020, an increasingly larger sector of the public expressed their belief that Jordan was moving in the negative direction, amid rising daily confirmed cases and high death rate, reaching 80.0% disapproval rate in late October 2020 [34]. These indications were in agreement with reports associating lower acceptance levels of COVID-19 vaccines with lower trust in government handling of the pandemic [28]. Such low acceptance levels should prompt the government to offer commensurate efforts in offering vaccine awareness campaigns that can regain people’s trust in their government handling of the ongoing crisis.

An important factor to consider when exploring vaccine acceptability is vaccine convenience in terms of its availability and affordability [20]. In the current study, the willingness to pay for the vaccine was a predictor of vaccine acceptance. According to the Jordanian Ministry of Health (MOH), the government will only be able to provide free COVID-19 vaccines for 20.0% of the population, while the rest will have to purchase it, without information whether it will be available at a subsidized price or not [35]. This should be factored in the government’s planning for vaccination programs and how acceptance level may change depending on the prices ascribed to the vaccines. In the current study, only 36.2% believed that the government will be able to provide the vaccine for free, indicating that the economic challenges faced by the Jordanian government may have played a role in shrinking the acceptance level. Further, the trust in the manufacturer that provides effective and noncontaminated products is another important determinant of confidence. About two-thirds of participants (59%) in the current study had confidence in pharmaceutical companies to develop safe and effective COVID-19 vaccines. However, the source of the vaccine affects the perceived safety, as only one-third of the participants in the current study perceived that COVID-19 vaccines that were manufactured in Europe or America were safer than those made in other countries. This is rather lower than the reported percentage by Pogue and colleagues where ∼55% and 36% of participants stated that they were more comfortable with vaccines made in the USA and Europe, respectively [33].

COVID-19 pandemic as with other previous pandemics is associated with feelings of fears, anxiety, and worries [36, 37]. However, it is unique in terms that people are not worried only about getting infected or transmit the disease to others [38], but they suffered societal and economic concerns due to the measures that were undertaken by the governments to confine the pandemic and stopping the human-human transmission of the disease [6]. These measures include enforcement of curfews and lockdowns (the largest throughout history), social distancing and self-isolation, schools and universities closures, borders’ shutdowns, travel restrictions, and quarantine [6]. In the current study, family members being infected (73.1%), topped the list of Jordan population worries during the COVID-19 pandemic followed by fears of death (30.0%), and then anxieties of becoming infected themselves (27.3%). The least-worries were financial related worries and being forced to take medication, respectively. Our findings were in alignment with the findings of Mertens et al, where increased fear during the COVID-19 pandemic was found to be related to perceived risks for family members and health anxiety [39]. Such a high percentage of fear over loved ones get infected could be attributed to the reports identifying elderly people with chronic diseases such as hypertension, diabetes, chronic respiratory disease, and weakened immune systems as a high-risk group to get infected with COVID-19 [40].

One of the themes of worries during the COVID-19 pandemic that were identified in a study from the Philippines is the worry of acquiring the diseases for self, family, and others [41]. Interestingly, the self-worry is focused mainly on preventing the transmission of the disease to family members especially older ones who were identified as vulnerable to COVID-19 [41].

Holingue et al showed in a population-based study of US adults that the fears and anxiety of getting infected with and die from COVID-19 were associated with increased mental distress [38]. Moreover, the personal hygienic precautions that were undertaken by individuals to avoid infecting others had increased the probability of becoming mentally distressed [38]. A systematic review and meta-analysis of the psychological and mental impact of COVID-19 showed that the prevalence of anxiety and depression was 33% and 28%, respectively [42].

During the COVID-19 pandemic, people used multiple information resources to gain knowledge and health information about the disease, including television, radio, newspapers, social media, friends, co-workers, healthcare providers, scientists, governments, etc [43]. Since such information sources can shape peoples’ acceptance or refusal of COVID-19 vaccines [44], it is crucial to disseminate transparent and accurate information about vaccines’ safety and efficacy to gain the trust of the population especially the hesitant and skeptical ones [45]. Hence, gaining an understanding of the resources that people trust the most to get information about COVID-19 vaccines is critical for the success of any future national vaccination campaign.

When Jordanians were asked about the most-trusted information sources about COVID-19 vaccines, health care providers topped the list, followed by pharmaceutical companies reports, and the national Jordanian government. The least-trusted information sources were social media and family members, respectively. Our findings are consistent with the KFF COVID-19 Vaccine Monitor: December 2020; where about 85% of the U.S. adults said they trust the most their doctors or healthcare providers for information related to COVID-19 vaccines, followed by national messengers (73%), and the FDA (70%) [46]. Our data were also in agreement with another study where the participants reported that they trust health care professionals the most, followed by their physicians [23]. In a further study, COVID-19 vaccine acceptance among college students in South Carolina was found to be affected by the information resources. Students largely trusted scientists (83%), followed by healthcare providers (74%), and then health agencies (70%) [47]. However, contrary to our study, college students do not trust information disseminated by pharmaceutical companies [47]. In a study from France, vaccination practices and acceptance toward MMR and HBV vaccines were better when parents had reported getting the information from their healthcare providers compared with parents getting information from the internet or their relatives [48]. The trust of the Jordanian population in the government as a source of information for COVID-19 vaccines is indicative of the peoples’ trust in the health system and the registration procedure of vaccines in Jordan.

## Conclusions

In conclusion, we identified Jordan as one of the lowest countries in the acceptance of COVID-19 vaccines, where a considerable percentage of the population of Jordan (36.3%) indicated a refusal to get vaccinated, while 26.3% were not sure. Vaccines perceived safety concerns and cost were associated with this refusal. Hence, the health authorities via health care providers, who were identified by the people as the most trust source of information regarding information about COVID-19 vaccines, should design interventions in terms of awareness campaigns via all types of multimedia to spread more transparent information about the safety and efficacy of the vaccines. The awareness campaigns should also shed the light over the new technology that was utilized in the production of few of them in order to boost COVID-19 vaccines acceptance. Making the vaccine available for free or at subsidized prices by the government could as well enhance vaccines acceptance among the population.

## Data Availability

Data used in this paper is available upon request.

## Acknowledgments

The financial support of the Deanship of Research, Jordan University of Science and Technology, Irbid, Jordan.

## References

1. World Health Organization. WHO Coronavirus Disease (COVID-19) Dashboard: World Health Organization; 2020 [cited 2020 13 December]. Available from: https://covid19.who.int/.

2. Helmy YA, Fawzy M, Elaswad A, Sobieh A, Kenney SP, Shehata AA. The COVID-19 Pandemic: A Comprehensive Review of Taxonomy, Genetics, Epidemiology, Diagnosis, Treatment, and Control. J Clin Med. 2020;9(4):1225. doi: 10.3390/jcm9041225. PubMed PMID: 32344679.

3. Wu F, Zhao S, Yu B, Chen Y-M, Wang W, Song Z-G, et al. A new coronavirus associated with human respiratory disease in China. Nature. 2020;579(7798):265–9. doi: 10.1038/s41586-020-2008-3.

4. Cucinotta D, Vanelli M. WHO Declares COVID-19 a Pandemic. Acta Biomedica. 2020;91(1):157–60. Epub 2020/03/20. doi: 10.23750/abm.v91i1.9397. PubMed PMID: 32191675; PubMed Central PMCID: PMCPMC7569573 stock ownership, equity interest, patent/licensing arrangement etc.) that might pose a conflict of interest in connection with the submitted article.

5. Phua J, Weng L, Ling L, Egi M, Lim C-M, Divatia JV, et al. Intensive care management of coronavirus disease 2019 (COVID-19): challenges and recommendations. Lancet Respir Med. 2020;8(5):506–17. doi: 10.1016/S2213-2600(20)30161-2.

6. Nicola M, Alsafi Z, Sohrabi C, Kerwan A, Al-Jabir A, Iosifidis C, et al. The socio-economic implications of the coronavirus pandemic (COVID-19): A review. International Journal of Surgery. 2020;78:185–93. doi: https://doi.org/10.1016/j.ijsu.2020.04.018.

7. Kaddoura M, AlIbrahim M, Hijazi G, Soudani N, Audi A, Alkalamouni H, et al. COVID-19 Therapeutic Options Under Investigation. Front Pharmacol. 2020;11:1196.

8. Beigel JH, Tomashek KM, Dodd LE, Mehta AK, Zingman BS, Kalil AC, et al. Remdesivir for the Treatment of Covid-19 — Final Report. New England Journal of Medicine. 2020;383(19):1813–26. doi: 10.1056/NEJMoa2007764.

9. FDA Approves First Treatment for COVID-19 [Internet]. 2020. Available from: https://www.fda.gov/news-events/press-announcements/fda-approves-first-treatment-covid-19

10. Rochwerg B, Agoritsas T, Lamontagne F, Leo Y-S, Macdonald H, Agarwal A, et al. A living WHO guideline on drugs for covid-19. BMJ. 2020;370:m3379. doi: 10.1136/bmj.m3379.

11. Hajj Hussein I, Chams N, Chams S, El Sayegh S, Badran R, Raad M, et al. Vaccines Through Centuries: Major Cornerstones of Global Health. Front Public Health. 2015;3:269.

12. Rodrigues CMC, Plotkin SA. Impact of Vaccines; Health, Economic and Social Perspectives. Front Microbiol. 2020;11:1526.

13. Ehreth J. The value of vaccination: a global perspective. Vaccine. 2003;21(27):4105–17. doi: https://doi.org/10.1016/S0264-410X(03)00377-3.

14. Zimmer C, Corum J, Wee S-L. Coronavirus vaccine tracker US: The New York Times; 2020 [cited 2020 December 13]. Available from: https://www.nytimes.com/interactive/.

15. Coustasse A, Kimble C, Maxik K. COVID-19 and Vaccine Hesitancy: A Challenge the United States Must Overcome. J Ambul Care Manage. 2021;44(1).

16. Omer SB, Yildirim I, Forman HP. Herd Immunity and Implications for SARS-CoV-2 Control. JAMA. 2020;324(20):2095–6. doi: 10.1001/jama.2020.20892.

17. Fine P, Eames K, Heymann DL. “Herd Immunity”: A Rough Guide. Clin Infect Dis. 2011;52(7):911–6. doi: 10.1093/cid/cir007.

18. Schoch-Spana M, Brunson EK, Long R, Ruth A, Ravi SJ, Trotochaud M, et al. The public’s role in COVID-19 vaccination: Human-centered recommendations to enhance pandemic vaccine awareness, access, and acceptance in the United States. Vaccine. 2020. doi: https://doi.org/10.1016/j.vaccine.2020.10.059.

19. MacDonald NE. Vaccine hesitancy: Definition, scope and determinants. Vaccine. 2015;33(34):4161–4. doi: https://doi.org/10.1016/j.vaccine.2015.04.036.

20. MacDonald NE. Vaccine hesitancy: Definition, scope and determinants. Vaccine. 2015;33(34):4161–4. Epub 2015/04/22. doi: 10.1016/j.vaccine.2015.04.036. PubMed PMID: 25896383.

21. Neergaard L, Fingerhut H. AP-NORC poll: Half of Americans would get a COVID-19 vaccine: Associated Press; May 28, 2020 [cited 2020 December 14]. Available from: https://apnews.com/article/dacdc8bc428dd4df6511bfa259cfec44.

22. Fisher KA, Bloomstone SJ, Walder J, Crawford S, Fouayzi H, Mazor KM. Attitudes Toward a Potential SARS-CoV-2 Vaccine: A Survey of U.S. Adults. Ann Intern Med. 2020. doi: 10.7326/M20-3569.

23. Malik AA, McFadden SM, Elharake J, Omer SB. Determinants of COVID-19 vaccine acceptance in the US. EClinicalMedicine. 2020;26:100495. Epub 2020/08/25. doi: 10.1016/j.eclinm.2020.100495. PubMed PMID: 32838242; PubMed Central PMCID: PMCPMC7423333.

24. Ministry of Health. COVID-19 Updates in Jordan: Ministry of Health; 2020 [cited 2020 13/12/2020]. Available from: https://corona.moh.gov.jo/en/MediaCenter/1491.

25. Jensehaugen J. Jordan and COVID-19: Effective Response at a High Cost. Oslo: PRIO, 2020.

26. Roya News English. Pfizer’s COVID-19 vaccine to be delivered to Jordan at end of January: Obeidat Amman: Roya News; 2020 [cited 2020 13/12/2020]. Available from: https://en.royanews.tv/news/23890/2020-12-03.

27. French J, Deshpande S, Evans W, Obregon R. Key Guidelines in Developing a Pre-Emptive COVID-19 Vaccination Uptake Promotion Strategy. Int J Environ Res Public Health. 2020;17(16). Epub 2020/08/23. doi: 10.3390/ijerph17165893. PubMed PMID: 32823775; PubMed Central PMCID: PMCPMC7459701.

28. Lazarus JV, Ratzan SC, Palayew A, Gostin LO, Larson HJ, Rabin K, et al. A global survey of potential acceptance of a COVID-19 vaccine. Nat Med. 2020:1–4. Epub 2020/10/22. doi: 10.1038/s41591-020-1124-9. PubMed PMID: 33082575; PubMed Central PMCID: PMCPMC7573523.

29. Al-Mohaithef M, Padhi BK. Determinants of COVID-19 Vaccine Acceptance in Saudi Arabia: A Web-Based National Survey. J Multidiscip Healthc. 2020;13:1657–63. Epub 2020/12/03. doi: 10.2147/JMDH.S276771. PubMed PMID: 33262600; PubMed Central PMCID: PMCPMC7686470.

30. Ababneh M, Jaber M, Rababa’h A, Ababneh F. Seasonal influenza vaccination among older adults in Jordan: prevalence, knowledge, and attitudes. Hum Vaccin Immunother. 2020;16(9):2252–6. doi: 10.1080/21645515.2020.1718438.

31. Abu-rish EY, Elayeh ER, Mousa LA, Butanji YK, Albsoul-Younes AM. Knowledge, awareness and practices towards seasonal influenza and its vaccine: implications for future vaccination campaigns in Jordan. Fam Pract. 2016;33(6):690–7. doi: 10.1093/fampra/cmw086.

32. Abu-Farha RK, Alzoubi KH, Khabour OF. Public Willingness to Participate in COVID-19 Vaccine Clinical Trials: A Study from Jordan. Patient Prefer Adherence. 2020;Volume 14:2451–8. doi: 10.2147/ppa.S284385.

33. Pogue K, Jensen JL, Stancil CK, Ferguson DG, Hughes SJ, Mello EJ, et al. Influences on Attitudes Regarding Potential COVID-19 Vaccination in the United States. Vaccines (Basel). 2020;8(4). Epub 2020/10/08. doi: 10.3390/vaccines8040582. PubMed PMID: 33022917; PubMed Central PMCID: PMCPMC7711655.

34. Center for Strategic Studies. Jordanian Index Surveys Series: Jordanian Street Pulse - 24 Amman: University of Jordan; 2020 [updated October 6; cited 2020 December 20]. Available from: http://jcss.org/ShowNewsAr.aspx?NewsId=862.

35. Jordan to Begin Covid-19 Vaccination Drive by February — Health Minister Amman: Jordan Times; 2020 [cited 2020 15 December]. Available from: https://www.jordantimes.com/news/local/govt-adamant-ensuring-transparency-covid-vaccine-procedures--%C2%A0media-minister.

36. Blakey SM, Abramowitz JS. Psychological Predictors of Health Anxiety in Response to the Zika Virus. J Clin Psychol Med Settings. 2017;24(3):270–8. doi: 10.1007/s10880-017-9514-y.

37. Wheaton MG, Abramowitz JS, Berman NC, Fabricant LE, Olatunji BO. Psychological Predictors of Anxiety in Response to the H1N1 (Swine Flu) Pandemic. Cognit Ther Res. 2012;36(3):210–8. doi: 10.1007/s10608-011-9353-3.

38. Holingue C, Kalb LG, Riehm KE, Bennett D, Kapteyn A, Veldhuis CB, et al. Mental Distress in the United States at the Beginning of the COVID-19 Pandemic. Am J Public Health. 2020;110(11):1628–34. doi: 10.2105/AJPH.2020.305857.

39. Mertens G, Gerritsen L, Duijndam S, Salemink E, Engelhard IM. Fear of the coronavirus (COVID-19): Predictors in an online study conducted in March 2020. J Anxiety Disord. 2020;74:102258. doi: https://doi.org/10.1016/j.janxdis.2020.102258.

40. Mueller AL, McNamara MS, Sinclair DA. Why does COVID-19 disproportionately affect older people? Aging. 2020;12(10):9959–81. doi: 10.18632/aging.103344.

41. Nicomedes CJC, Avila RMA. An analysis on the panic during COVID-19 pandemic through an online form. J Affect Disord. 2020;276:14–22. doi: https://doi.org/10.1016/j.jad.2020.06.046.

42. Luo M, Guo L, Yu M, Jiang W, Wang H. The psychological and mental impact of coronavirus disease 2019 (COVID-19) on medical staff and general public – A systematic review and meta-analysis. Psychiatry Res. 2020;291:113190. doi: https://doi.org/10.1016/j.psychres.2020.113190.

43. Ali SH, Foreman J, Tozan Y, Capasso A, Jones AM, DiClemente RJ. Trends and Predictors of COVID-19 Information Sources and Their Relationship With Knowledge and Beliefs Related to the Pandemic: Nationwide Cross-Sectional Study. JMIR Public Health Surveill. 2020;6(4):e21071. doi: 10.2196/21071.

44. Freed GL, Clark SJ, Butchart AT, Singer DC, Davis MM. Sources and Perceived Credibility of Vaccine-Safety Information for Parents. Pediatrics. 2011;127(Supplement 1):S107. doi: 10.1542/peds.2010-1722P.

45. Siegrist M, Zingg A. The role of public trust during pandemics: Implications for crisis communication. Eur Psychol. 2014;19(1):23–32. doi: 10.1027/1016-9040/a000169.

46. Hamel L, Kirzinger A, Muñana C, Brodie M. KFF COVID-19 Vaccine Monitor: December 2020. KFF Health Tracking Poll Dec 15, 2020.

47. Qiao S, Friedman DB, Tam CC, Zeng C, Li X. Vaccine acceptance among college students in South Carolina: Do information sources and trust in information make a difference? medRxiv. 2020:2020.12.02.20242982. doi: 10.1101/2020.12.02.20242982.

48. Charron J, Gautier A, Jestin C. Influence of information sources on vaccine hesitancy and practices. Med Mal Infect. 2020;50(8):727–33. doi: https://doi.org/10.1016/j.medmal.2020.01.010.

